# Digital technology as a tool to provide social support to individuals with cancer in low- and middle-income countries: a scoping review

**DOI:** 10.1101/2024.08.09.24311634

**Authors:** Hallie Dau, Fazila Kassam, Beth A. Payne, Hana Miller, Gina Ogilvie

## Abstract

**BACKGROUND:** Cancer is a rising cause of morbidity and mortality in low- and middle-income countries (LMICs). Individuals diagnosed with cancer in LMICs often have limited access to cancer prevention, diagnosis, and treatment services. Social support after a cancer diagnosis is associated with positive health outcomes in the long term. Digital technologies, such as the Internet and mobile phones, could be used to provide support to individuals with cancer in a more accessible way. This scoping review aims to understand how digital technology is currently being utilized by individuals with cancer for social support in LMICs.

**METHODS:** Four electronic databases were searched up to June 2024 to identify studies that reported on the use of digital technology for cancer social support in LMICs. Articles were included if they were published in English, included adults diagnosed with any type of cancer, and reported the use of digital technology for social support. Study characteristics, population demographics, and technological interventions reported were extracted.

**RESULTS:** In all, 15 articles from 12 studies were included in the scoping review. The results were centralized in four countries, and the most common cancer type reported was breast. Online health communities, Internet-based resources, mobile applications, and telecommunication were the four digital technologies reported. Overall, the articles demonstrated that the use of digital technology for social support can be beneficial for individuals diagnosed with cancer in LMICs.

**CONCLUSION:** There is a limited understanding of how digital technology can be used to support individuals with cancer in LMICs. Future research is needed to explore how digital technology can be utilized by underrepresented regions to offer avenues of support for regionally common cancer types such as cervical. Fundamentally, this scoping review highlights the need for additional research on the use of digital technology to support individuals with a cancer diagnosis in LMICs.

## INTRODUCTION

Cancer is rising within low- and middle-income countries (LMICs) with the number of new cases annually in LMICs rapidly increasing. By 2030, it is anticipated that three-quarters of global cancer-related deaths will occur in LMICs.(1) In 2022 the Lancet Oncology Commission published a series highlighting the urgent need to focus on the increasing rates of cancer in Africa, an area that has been historically understudied.(2) The increase in cancer rates in Africa and similar regions can be attributed to urbanization, increased exposure to carcinogenic risk factors, an aging population, sedentary lifestyles, and limited access to cancer prevention, diagnosis, and treatment services.(2, 3) The burden of cancer in LMICs is a major public health challenge that requires urgent attention and action. Efforts to address this challenge should focus on improving cancer prevention, early detection, access to quality care, and risk factor reduction.

An opportunity to increase the quality of cancer care lies in sources of global connectivity. As of 2022, 82% of people in low-middle income countries have access to and use the internet.(4) More importantly, three-quarters of the world’s population owns a mobile phone - 49% of people in low-income countries and 65% of individuals in lower-middle-income countries.(4) With this, the rise of social media has allowed individuals to connect more easily with one another across the globe. As of 2022, Facebook (2.96 billion users) is the most popular social network followed by YouTube (2.51 billion users) and WhatsApp (2 billion users).(5) The opportunities for connection and healthcare access offered by technology should be leveraged to increase access to essential health services such as cancer support. The utilization of technology for cancer support is aligned with the World Health Organization’s prioritization of self-care interventions. As there is currently a global healthcare worker shortage, self-care interventions, particularly those based in digital technology, allow individuals to actively engage in advancing their own health and well-being in a convenient and cost-effective manner that reduces the strain on the healthcare system.(6)

Prior research has demonstrated that social support soon after a cancer diagnosis is associated with favorable health outcomes for individuals with cancer in the long term.(7, 8) Additionally, social support positively influences a patient’s help-seeking behavior.(9) Unfortunately, in many LMICs, social support is often lacking due to stigma related to diagnosis, poor health system infrastructure, and gender discrimination.(9) Digital technologies such as mobile phones and the internet offer the opportunity for cancer patients in LMICs to receive social support in a more accessible way. Prior research has demonstrated that online cancer communities can decrease psychological distress,(10, 11) improve support,(12, 13) and create feelings of empowerment.(14-16) To date, there is a large scale of evidence on the benefits of digital support for individuals with cancer.(17-20) However, there is limited knowledge about how social support interventions could be applied to individuals with cancer in LMICs. It is important to better understand how digital technologies can improve access to social support and ultimately improve cancer outcomes in LMICs. As the rates of cancer continue to rise in LMICs, the urgent need to address the lack of comprehensive cancer social support programs is amplified. The goal of this scoping review is to understand how digital technology is currently being utilized by individuals with cancer for social support in LMICs.

## METHODS

### Inclusion & exclusion criteria

Articles were included in the review if they (i) were published in English; (ii) were conducted in a LMIC (as defined by to the World Bank criteria);(21) (iv) included adults (≥18 years of age) diagnosed with any type of cancer; and (v) reported the use of digital technology for cancer social support.

Articles were excluded from the review if they (i) focused solely on children; and (ii) were review articles.

### Search strategy & extraction

The electronic databases, MEDLINE, PsychInfo, CINAHL, and CABI Global Health were searched up to June 2024 to identify eligible studies. Search terms are provided in **Supplementary 1**. Covidence systematic review software(22) was used to screen and extract all data from the included studies. Two reviewers independently screened studies and one author extracted data from included studies. Studies were first screened by title and abstract, subsequently, included studies underwent full-text review. The relevant data was extracted from the final included studies. Disagreements were discussed by the study team and resolved by reaching consensus.

The study population characteristics extracted by the authors included age, gender, education status, employment status, marital status, income, cancer stage, cancer type, and treatment type, when applicable. In addition, the type of digital technology utilized was also extracted. Following extraction, the data was grouped by the type of digital technology reported. The digital technology was compared and contrasted with regards to its usability and reported outcomes.

Social support was based on Cooke et al. and defined as “any emotional, informational, instrumental or appraisal support given to cancer survivors or patients to help them cope with biological, psychological or social stressors.”(23) The definition for digital health technology is based on the World Health Organization’s definition for digital health which defines it as “the use of information and communications technology in support of health and health-related fields.”(24) Digital health technology includes web-based resources, mobile applications, social media platforms, general internet use, and telecommunication.

## RESULTS

In all, 7848 original articles were identified through the search strategy and 15 were included in the final review (**Figure 1**). Three articles were from the same larger study and were combined into one study, resulting in 12 unique studies.(25-27) **Table 1** summarizes the characteristics of the included studies. The majority of included studies were either cross-sectional(28-33) (n=6) in design or a randomized control trial(25-27, 34-36) (n=4). All but one article(26) used quantitative methods. Among the 12 unique studies included in this review, only four countries were studied: China(37) (25-27, 29, 32-34, 36, 38) (n=8), Iran(30, 35) (n=2), Kenya(31) (n=1), and Serbia(28) (n=1). Breast(25-27, 29, 31, 37, 38) (n=5) was the most common cancer studied.

**Figure 1.**
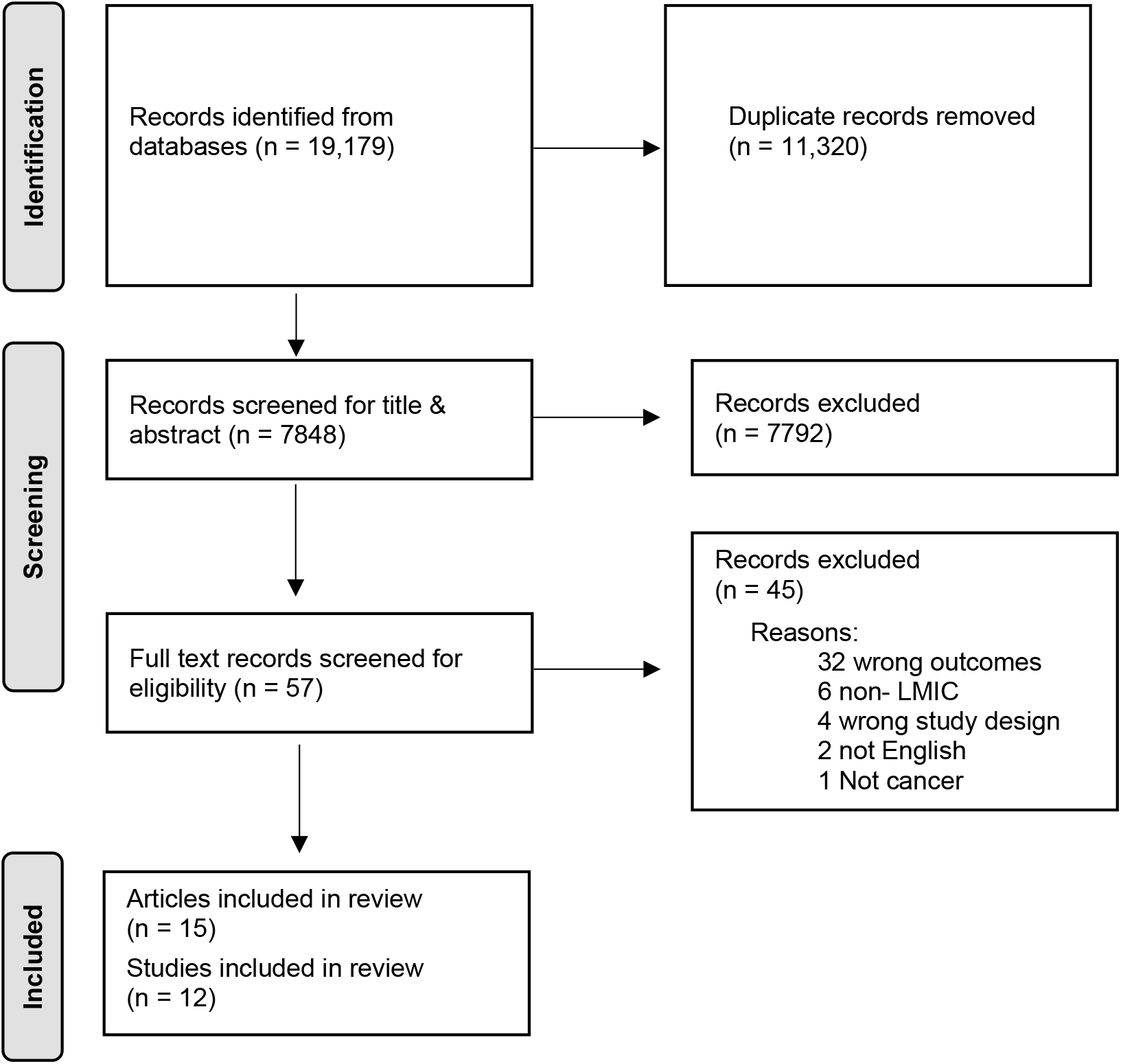
PRISMA flow diagram

**Table 1.** Characteristics of included studies.

| **Author** | **Year** | **Country** | **Study Type** | **Sample Size** | **Cancer Type** | **Digital technology utilized** |
| --- | --- | --- | --- | --- | --- | --- |
| Chen et al.(37) | 2024 | China | Pre-test-post-test | 20 | Breast | Mobile application |
| Klikovac(28) | 2015 | Serbia | Cross-sectional | 2,748 | Multiple cancers | Telephone |
| Lei et al.(29) | 2021 | China | Cross-sectional | Not reported | Lung, Breast | Online health community |
| Lin et al.(34) | 2023 | China | Randomized control trial | 168 | Gynecologic | Mobile application |
| Rahimi et al.(35) | 2021 | Iran | Randomized control trial | 60 | Colorectal | Telephone and virtual social networks |
| Salajegheh et al.(30) | 2020 | Iran | Cross-sectional | 350 | Multiple cancers | Internet, social networks, telephone, satellite, radio |
| Shaikh et al.(31) | 2021 | Kenya | Cross-sectional | 114 | Breast | Internet |
| Wang(36) | 2022 | China | Randomized control trial | 72 | Leukemia | Mobile application |
| Wang(32) | 2023 | China | Cross-sectional | 425 | Pancreatic | Internet, social networks |
| Wu et al.(38) | 2020 | China | Quasi-experimental | 60 | Breast | Mobile application |
| Zhou et al.(33) | 2020 | China | Cross-sectional | 162 | Multiple cancers | Online health community |
| Zhu et al.(25-27) | 2018 | China | Qualitative,<br>Randomized control trial | 13 (qualitative)<br>114 (quantitative) | Breast | Mobile application |

There were four primary types of digital technologies reported for cancer social support in the 15 included studies: online health communities, Internet-based resources, mobile applications, and telecommunication.

Cross-sectional studies by Zhou et al. and Lei et al. investigated the role of online health communities (OHCs). Zhou et al. conducted a cross-sectional survey among 162 participants engaged in OHCs in China. They found that e-health literacy was positively associated with the informational and emotional support a cancer survivor received by actively interacting in the OHC.(33) Lei et al. (2021) conducted a social network analysis of user activity and user-generated content in three forums and found that while a few members of the community are very well connected with many other members, most other members have fewer connections. Salient topics across all three forums were related to disease treatment, examination, diagnosis, social life, and healthy behavior change.

Three studies reported on Internet-based resources for cancer social support. The first, Shaikh et al., developed a website to address the unmet needs, including social support, for women with metastatic breast cancer in Kenya.(31) Over a two-year period, the website received 7,864 unique visitors. Among the 114 women with metastatic breast cancer they interviewed during their needs assessment, 90.4% had moderate to high supportive care needs, with psychological being the highest (63%). Given the high need for social support, the authors concluded that interactive Internet-based resources could be useful for women living with metastatic breast cancer in countries with limited health services.(31) Salajegheh et al.(30) and Wang(32) were both cross-sectional studies that surveyed individuals with cancer about their Internet usage. Salajegheh et al. reported that among the 350 individuals with cancer that they surveyed, the Internet was one of the least likely ways individuals sought out information.(30) Wang surveyed pancreatic cancer internet users and found that social media use has a positive impact on mental health, namely anxiety and depression.(32)

Five unique studies, all conducted in China, reported on the use of mobile applications for cancer support. (26, 27, 34, 36, 38, 39) Chen et al, Wang et al., and Wu et al. evaluated ways WeChat, a Chinese social media and messaging application, could be used to help support individuals with cancer in China.(36-38) All three interventions provided information and support through the mobile application. Wang et al. included the WeChat platform as well as a micro-website developed by the researchers,(36) Wu et al. recruited nurses to lead and interact with participants through the platform, (38) and Chen et al. developed an interactive platform.(37) At the end of the study period, both Wang et al. and Wu et al. reported that those in the WeChat group had higher psychological well-being than those in the control.(36, 38) Additionally, Wang et al. and Chen et al. reported improved quality of life by the end of the study period.(36, 37) For example, in Wang et al. the quality of life score (range 0-100) improved from 51.1 in the pre-intervention to 63.1 in the post-intervention.(36) Wu et al. reported that those who received the intervention reported a higher level of social support outside of family.(38)

Both Zhu et al. (25-27) and Lin et al.(34) measured participant’s self-efficacy after using a mobile application created by the study team. Both applications include a discussion forum for participants to engage in. Zhu et al. found that the program improved emotional well-being,(26) self-efficacy,(27) quality of life, (27) and social support among individuals with breast cancer. (27) Lin et al. found that individuals with gynecological cancer in the intervention group had lower levels of illness uncertainty and higher quality of life levels.(34)

Two studies reported on the use of telecommunications.(28, 30, 35) Klikovac established a telephone service to provide psychological support to cancer patients of all types and stages, as well as their family members in Serbia. They found that most of the calls were regarding emotional support for sadness, hopelessness, depression and informational support for disease prognosis, treatment, and side effects of treatment.(28) Rahimi et al. conducted a double-blind randomized control trial to explore how peer support via telephone and social networks impacts colorectal cancer patients. They reported that participants who received informational, emotional, and appraisal support from their peers via telephone two times a week and virtual social networks three times a week had overall higher well-being than the control group.(35)

## DISCUSSION

In all, 15 articles and 12 studies were included in this scoping review. The majority of the studies were conducted in China. Online health communities, Internet-based resources, mobile applications, and telecommunication were the four primary types of digital technology utilized for cancer social support. Overall, the review found that the use of digital technology for social support can be beneficial for individuals diagnosed with cancer in LMICs. More specifically, we found that digital technology can improve quality of life, reduce anxiety and depression, and allow individuals to connect with other individuals diagnosed with cancer.

Only four countries were represented in this review. The majority of the included studies were from China,(37) (25-27, 29, 32-34, 36, 38) a country that has the most internet users worldwide,(40) as well as the second highest gross-domestic product globally.(41) There was only one study from the African region (Kenya)(31) and no research from South America and Central Asia. This likely is because cancer and other non-communicable diseases have largely been understudied in LMICs with the primary focus being on communicable diseases.(2, 42) As such, there is a gap in understanding how digital technology can be used effectively to provide social support for cancer patients across the broad category of LMICs. It is important to study this topic more widely since many LMICs are similar in that they lack robust healthcare systems that can adequately care for all the physical and social needs of individuals diagnosed with cancer. (1, 3, 43) More widespread and varied research on digital platforms can better lead to the adaptation and adoption of programs across more resource-limited countries. Social support through digital technology could reduce the overall burden of care on the local healthcare system.

Similar to a deficiency in the regions studied, there is a limited understanding of how digital technology can support various cancer types. The majority of the included studies reviewed breast cancer. However, the top five types of cancer in LMICs are breast, cervical, liver, colorectal, and lip/oral.(44) Given these cancers differ in their treatment of care, the way in which technology can be used for social support may differ as well. Evidently, cancers associated with significant morbidity and mortality in LMICs such as cervical(45) have not been the focus of digital health interventions and thus, current evidence may not be applicable to all cancer patients from these regions. Therefore, there needs to be a greater understanding of how digital technology can best provide social support for not only a variety of cancer types, but those most common to LMICs.

The results of this review demonstrate that despite the surging cases of cancer in LMICs, there has been limited exploration of the opportunities to provide social support to individuals with cancer using digital technologies. More specifically, there is a need to better understand how existing and commonly utilized technological interventions can be implemented for cancer social support in LMICs. To date, the majority of research has focused on mobile applications followed by general Internet use. This is only a small fraction of what could be harnessed to support individuals with cancer in LMICs. For example, with regards to social media, WeChat is the only platform that has been studied for digital social support for individuals with cancer in LMICs.(38) However, WeChat is designed primarily for Chinese users and as such the findings are not applicable to other countries. Consequently, there is an urgent need for research on how the more commonly used and growing platforms like WhatsApp, Facebook, and YouTube can be adapted for digital social support for cancer patients in LMICs. For example, research from the United States and Australia has shown that social media can provide positive social support benefits for individuals diagnosed with cancer(46, 47) and that there is an interest in it, particularly among young adults with cancer.(47-49) Replicating these findings in an LMIC setting should be a research priority.

This scoping review was strengthened by its comprehensive search strategy that, in turn, identified multiple gaps in relation to the use of digital technology for social support for individuals diagnosed with cancer in LMICs. Additionally, the review adhered to best practices to reduce bias in results and included two authors to review potential articles. This review was limited in that the authors only searched for studies published in English. As such, publication bias may have occurred.

Furthermore, the authors did not consider unpublished or grey literature which may have provided additional results.

Despite the vast advancements in technology, there is limited evidence for its use to deliver social support to cancer patients in LMICs. Only 12 studies were identified in this scoping review, and they were limited to four countries, the majority being China. Future research should consider including more nations in underrepresented regions, cancer types that carry the greatest burden in LMICs, and widely used and acceptable digital platforms. Furthermore, longitudinal studies are needed to determine how long the benefits of digital technology last after a diagnosis. As individuals continue to spend increasing amounts of time in the digital space, it is important for the cancer community to focus their attention on harnessing this technology for better support and overall care. This in turn will help reduce the burden of care on local healthcare systems and lead to more positive health outcomes for individuals diagnosed.

## Data Availability

All data produced in the present work are contained in the manuscript

