## Supplementary 1 for "Digital technology as a tool to provide social support to individuals with cancer in low- and middle-income countries: a scoping review"

**Search Strategy**

internet OR cyber OR web-based OR “web based” OR online OR computer OR tablet OR ipad OR desktop OR laptop OR “social medium” OR multimedia OR “social media” OR twitter OR facebook OR instagram OR twitter OR pinterest OR podcast OR blog OR forum OR “social network”* OR blog* OR vlog OR “video blog” OR youtube OR microblog OR zoom OR skype OR facetime OR “video chat” OR whatsapp OR wechat OR tumblr OR “Sina Weibo” OR qq OR Kuaishou OR qzone OR “microsoft teams” OR discord OR google OR messenger OR imjur OR vimeo OR tiktok OR quora OR clubhouse OR 4chan OR viber OR snapchat OR “chat room”* OR linkedin OR telegram OR reddit OR podcast* OR wordpress OR “square space” OR phone OR telephone OR cellphone OR wireless OR mobile OR cellular OR smartphone OR “text messag”* OR texting OR mms OR sms OR “instant messag”* OR bbm OR telemedicine OR “telephone medicine” OR ehealth OR “e health” OR “electronic health” OR telenursing OR “telephone nursing” OR mhealth OR “mobile health” OR app OR application OR apps

AND

Cancer* OR neoplasm* OR carcinoma* OR adenocarcinoma* OR malig* OR tumo?r* OR metasta* OR oncolog* OR sarcoma*

AND

Support* OR connect* OR communicat* OR stigma* OR isolat* OR social* OR recover* OR “side effect”* OR question* OR chat* OR talk* OR peer OR network* OR friend* OR psychosocial OR listen* OR companion OR belong* OR self-help OR “self help” OR “help group” OR “patient group”

AND

afghanistan OR albania OR algeria OR “american samoa” OR angola OR argentina OR armenia OR armenian OR azerbaijan OR bangladesh OR “republic of Belarus” OR belarus OR byelarus OR belorussia OR byelorussian OR belize OR “british honduras” OR benin OR bhutan OR bolivia OR "bosnia and herzegovina" OR bosnia OR herzegovina OR botswana OR brazil OR brasil OR bulgaria OR “burkina faso” OR “burkina fasso” OR burundi OR “cabo verde” OR cambodia OR cameroon OR cameron OR cameroun OR “central African republic” OR chad OR china OR colombia OR comoros OR “democratic republic of the congo” OR “democratic republic congo” OR congo OR “costa rica” OR "cote d’ivoire" OR "cote d’ ivoire" OR “cote divoire” OR “cote d ivoire” OR “ivory coast” OR cuba OR djibouti OR “dominican republic” OR ecuador OR egypt OR “el Salvador” OR “equatorial guinea” OR eritrea OR eswatini OR ethiopia OR fiji OR gabon OR gambia OR "georgia (republic)" OR ghana OR grenada OR guatemala OR guinea OR “guinea Bissau” OR guyana OR haiti OR honduras OR india OR indonesia OR iran OR iraq OR jamaica OR jordan OR kazakhstan OR kenya OR "democratic people’s republic of korea" OR “north korea” OR kosovo OR kyrgyzstan OR “kyrgyz republic” OR laos OR “lao pdr” OR "lao people's democratic republic" OR lebanon OR “lebanese republic” OR lesotho OR liberia OR libya OR "macedonia (republic)" OR macedonia OR madagascar OR malaysia OR maldives OR mali OR micronesia OR “federated states of Micronesia” OR kiribati OR “marshall islands” OR tuvalu OR mauritania OR mexico OR moldova OR mongolia OR montenegro OR morocco OR mozambique OR myanmar OR burma OR namibia OR nepal OR nicaragua OR niger OR nigeria OR pakistan OR “papua new guinea” OR “new guinea” OR paraguay OR peru OR philippines OR philipines OR phillipines OR phillippines OR russia OR “russian federation” OR rwanda OR ruanda OR samoa OR "sao tome and principe" OR senegal OR serbia OR “solomon island” OR “solomon islands” OR Somalia OR “south Africa” OR “south sudan” OR “sri lanka” OR “saint lucia” OR "st. lucia" OR "saint vincent and the grenadines" OR “saint Vincent” OR "st. vincent" OR “grenadines” OR sudan OR suriname OR Surinam OR Syria OR “syrian arab republic” OR Tajikistan OR tadjikistan OR Tadzhikistan OR tanzania OR tanganyika OR Thailand OR “timor leste” OR “east timor” OR togo OR “togolese republic” OR tonga OR tunisia OR turkey OR "turkey (republic)" OR Turkmenistan OR turkmen OR Uganda OR ukraine OR uzbekistan OR uzbek OR Vanuatu OR Venezuela OR Vietnam OR “viet nam” OR “middle east” OR “west bank” OR gaza OR yemen OR Zambia OR Zimbabwe OR “global south” OR “low income countr”* OR “low and middle income country” OR “low and middle income countries” OR LMIC*
